# Sero-monitoring of health care workers reveals complex relationships between common coronavirus antibodies and SARS-CoV-2 severity

**DOI:** 10.1101/2021.04.12.21255324

**Authors:** Sigrid Gouma, Madison E. Weirick, Marcus J. Bolton, Claudia P. Arevalo, Eileen C. Goodwin, Elizabeth M. Anderson, Christopher M. McAllister, Shannon R. Christensen, Debora Dunbar, Danielle Fiore, Amanda Brock, JoEllen Weaver, John Millar, Stephanie DerOhannessian, The UPenn COVID Processing Unit, Ian Frank, Daniel J. Rader, E. John Wherry, Scott E. Hensley

## Abstract

Recent common coronavirus (CCV) infections are associated with reduced COVID-19 severity upon SARS-CoV-2 infection, however the immunological mechanisms involved are unknown. We completed serological assays using samples collected from health care workers to identify antibody types associated with SARS-CoV-2 protection and COVID-19 severity. Rare SARS-CoV-2 cross-reactive antibodies elicited by past CCV infections were not associated with protection; however, the duration of symptoms following SARS-CoV-2 infections was significantly reduced in individuals with higher common betacoronavirus (βCoV) antibody titers. Since antibody titers decline over time after CCV infections, individuals in our cohort with higher βCoV antibody titers were more likely recently infected with common βCoVs compared to individuals with lower antibody titers. Therefore, our data suggest that recent βCoV infections potentially limit the severity of SARS-CoV-2 infections through mechanisms that do not involve cross-reactive antibodies. Our data are consistent with the emerging hypothesis that cellular immune responses elicited by recent common βCoV infections transiently reduce disease severity following SARS-CoV-2 infections.

## Introduction

SARS-CoV-2 causes heterogenous disease outcomes in different individuals that can range from asymptomatic infections to critical illness and death (1). It is unknown if prior exposure histories to common coronaviruses (CCVs) contribute to diverse outcomes following SARS-CoV-2 infections. A study reviewing electronic health records indicated that individuals recently infected with CCVs were not protected from SARS-CoV-2 infections but experienced less severe disease upon infection (2). Our group and others have found that some individuals possessed pre-pandemic antibodies that cross-react to SARS-CoV-2 (3-5), but these cross-reactive antibodies were not associated with SARS-CoV-2 protection or attenuating COVID-19 severity. Thus, it is unclear how prior CCV exposures influence outcomes following SARS-CoV-2 infections.

Antibody titers to CCVs are elevated after recent CCV infections but then gradually decline over time (6). Antibody titers to CCVs can therefore serve as an ‘immunological stamp’ that dates recent CVV infections. Much less is known about the kinetics of T cell responses following CCV infections and how cellular immunity elicited by past CCV exposures impacts subsequent encounters with CCVs and SARS-CoV-2. Some individuals possessed SARS-CoV-2-reactive CD4+ and CD8+ T cells prior to the COVID-19 pandemic (7-11); however, the impact of cellular immunity elicited by prior common coronavirus infections on SARS-CoV-2 infections is poorly understood.

In this study we established a cohort of 2,043 health care workers and we longitudinally collected serum samples in the spring and summer of 2020 during the first wave of SARS-CoV-2 activity in Philadelphia, PA. We identified a subset of health care workers who went on to become infected with SARS-CoV-2 after we collected serum samples. We completed a series of serological assays to determine if antibodies reactive to SARS-CoV-2 and CCVs were associated with SARS-CoV-2 protection and COVID-19 severity upon infection.

## Results

### Establishment of a health care worker cohort

We established a prospective cohort of 2,043 health care workers during the spring of 2020 to monitor SARS-CoV-2 seroprevalence and identify correlates of protection against SARS-CoV-2 infections. We included health care workers at 3 hospitals in the University of Pennsylvania health system (Hospital of the University of Pennsylvania, Penn Presbyterian Medical Center, and Pennsylvania Hospital) who had direct contact with or worked on units with patients, and we excluded anyone previously diagnosed with COVID-19. Participants were predominantly female (75.2%), White (82.9%) and non-Hispanic (96.5%). The median age was 36 years (inter quartile range [IQR]: 30-46 years) (**Supplemental Table 1**). Participants of our study were enrolled during the spring of 2020 when SARS-CoV-2 began widely circulating in Philadelphia (**Figure 1A**).

**Figure 1.**
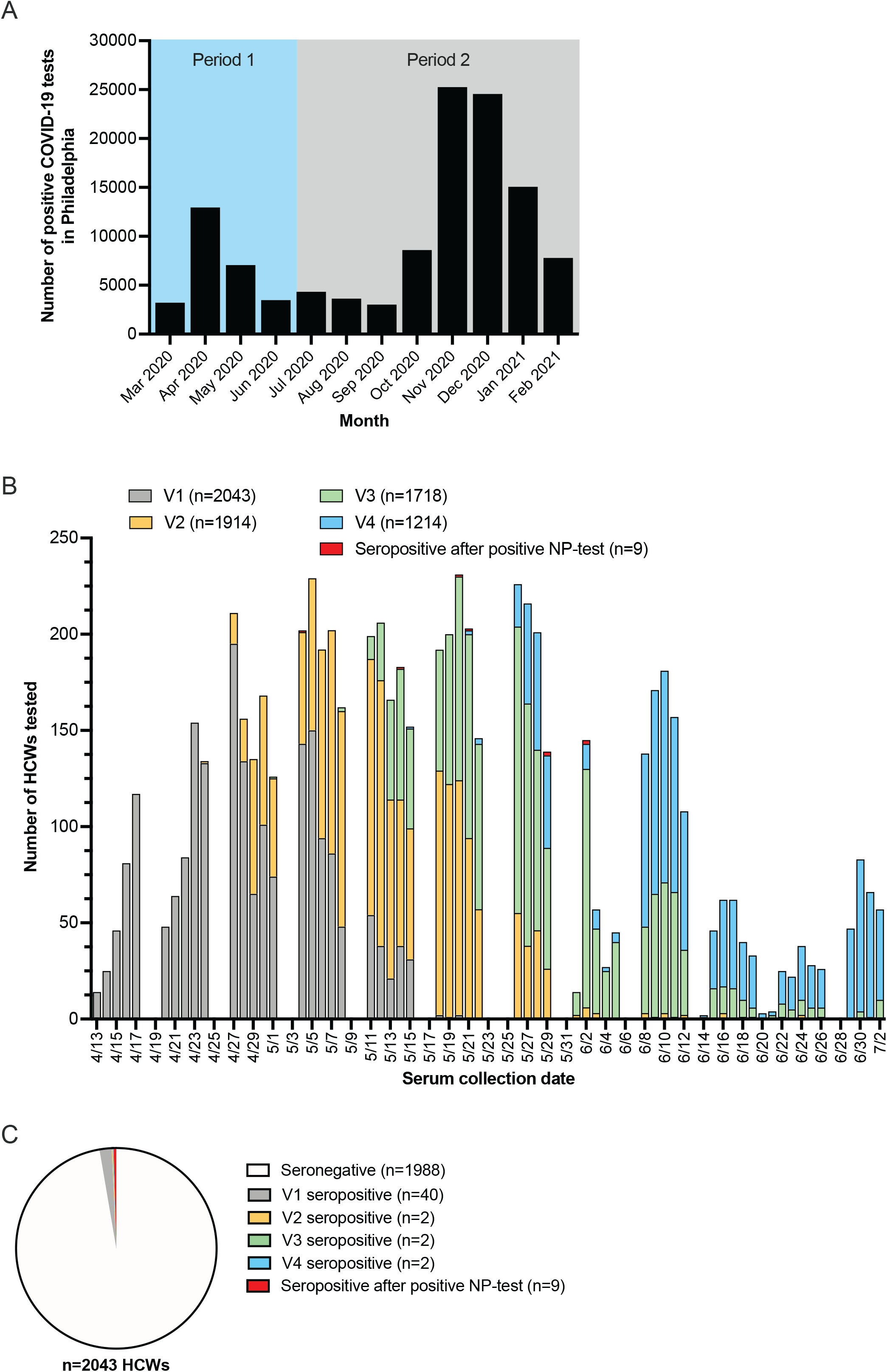
Seropositive health care workers by study visit in relation to SARS-CoV-circulation in Philadelphia. (A) Number of positive COVID-19 tests in Philadelphia from March 2020 – February 2021 (data retrieved from opendataphilly.org on 9 March 2021). The first viral period is defined as infections that occurred before July 2, 2020 and the second viral period as infections that occurred after July 2, 2020. (B) Number of health care workers tested by serum collection date, stratified by study visit and seropositivity status. One out of the 9 health care workers with a positive NP SARS-CoV-2 PCR test outside of our study seroconverted after 2 July 2020 and their seropositive sample is therefore not shown in this graph. (C) Seropositive health care workers (n=55) by study visit. The majority of health care workers (n=1988) were seronegative throughout the study period.

We collected baseline serum samples from each participant between April 13, 2020 and May 20, 2020 (**Figure 1B**). Within 36-48 hours after sample collection, we quantified levels of SARS-CoV-2 spike receptor binding domain (S-RBD) serum antibodies. We collected NP swabs from all SARS-CoV-2 S-RBD seropositive participants and we completed SARS-CoV-2 PCR testing to identify active or recent infections. Participants who were seronegative at the baseline visit were invited for follow-up visits every 2 weeks until July 2, 2020 and NP SARS-CoV-2 PCR testing was completed on all participants who seroconverted. In total, we collected 6,897 serum samples between April 13, 2020 and July 2, 2020 from 2,043 health care workers (**Figure 1B**). This included 2,043 serum samples collected at a baseline visit, 1,914 samples collected at visit 2, 1,718 samples collected at visit 3, and 1,214 samples collected at visit 4. We also collected serum samples from 8 participants who were seronegative at their baseline visit and had a positive NP SARS-CoV-2 PCR test outside of our study before July 20, 2020. Additional serum samples were collected from seropositive health care workers up to 236 days post-seroconversion to monitor the longevity of antibody responses following SARS-CoV-2 infection, including one seropositive participant who had a positive NP SARS-CoV-2 PCR after July 2, 2020.

### Seroprevalence during the first spring/summer 2020 wave of SARS-CoV-2 infections in Philadelphia

We found that 40 of 2,043 healthcare workers (2.0%) in our cohort possessed serum IgG and/or IgM SARS-CoV-2 S-RBD-reactive antibodies at baseline (**Figure 1C**). Of the 40 seropositive samples, 17 (42.5%) were IgG^+^/IgM^-^, 7 (17.5%) were IgG^-^/IgM^+^, and 16 (40.0%) were IgG^+^/IgM^+^. Seropositivity remained consistently low throughout the study period (**Figure 1B**). Out of the 2,003 health care workers who were seronegative at baseline, 15 health care workers (0.7%) became seropositive on subsequent study visits (**Figure 1C**). Of the 15 health care workers who seroconverted while enrolled in our study, 5 (33.3%) were IgG^+^/IgM^-^ and 10 (66.7%) were IgG^+^/IgM^+^. As of July 2, 2020, the overall seropositivity rate in our health care workers cohort was 2.7%, which was similar to the 3.2% seroprevalence rate reported for the Philadelphia metro area and surrounding counties from 13-25 April 2020 (12), but lower than the 6.2% seroprevalence rate we previously reported in samples collected from parturient women from 4 April-3 June 2020 (13). Consistent with other reports (14, 15), the low seroprevalence within our cohort suggest that PPE and other precautions taken within our hospitals limited the spread of SARS-CoV-2 infections of healthcare workers.

Of the 40 health care workers who were seropositive at baseline, only 6 (15.0%) were NP SARS-CoV-2 PCR positive. Since we only enrolled health care workers who did not have a known or suspected history of COVID-19 diagnosis, most seropositive participants entering our study likely had prior asymptomatic SARS-CoV-2 infections before the study began. Of the 15 health care workers who seroconverted after the baseline visit, 12 (80.0%) were SARS-CoV-2 NP PCR positive.

### Antibody kinetics after SARS-CoV-2 infection

We stopped collecting blood samples from seronegative participants in July of 2020 after the first SARS-CoV-2 wave in Philadelphia; however, we continued to collect samples from seropositive health care workers so that we could measure serum antibody levels within infected individuals over time. We quantified SARS-CoV-2 S-RBD antibody levels in samples longitudinally collected from 47 seropositive participants who possessed IgG antibodies, including 33 health care workers who possessed IgG antibodies to SARS-CoV-2 S-RBD at the first study visit and 14 health care workers who seroconverted after first sample collection (**Figure 2**). Consistent with previous reports (16-19), SARS-CoV-2 S-RBD IgG levels remained relatively stable in the serum of the majority of health care workers (**Figure 2A**). Of the 39 health care workers with samples collected for 140 days post seroconversion, 33 possessed detectable levels of serum IgG antibodies. Health care workers with undetectable IgG concentrations at 140 days post seroconversion had significantly lower peak geometric mean IgG concentrations than health care workers who possessed IgG antibodies at 140 days post seroconversion (0.66 versus 6.35 arbitrary units/mL; p<0.001 in unpaired t test on log_2_-transformed data). In contrast to IgG levels, the longevity of the IgM antibody response was highly variable among participants (**Figure 2B**). We detected serum IgM in some participants for multiple weeks, including one participant with detectable IgM up to 168 days post seroconversion. Interestingly, 5 out of 14 participants who seroconverted did not possess detectable serum IgM at any study visit (**Figure 2B**).

**Figure 2.**
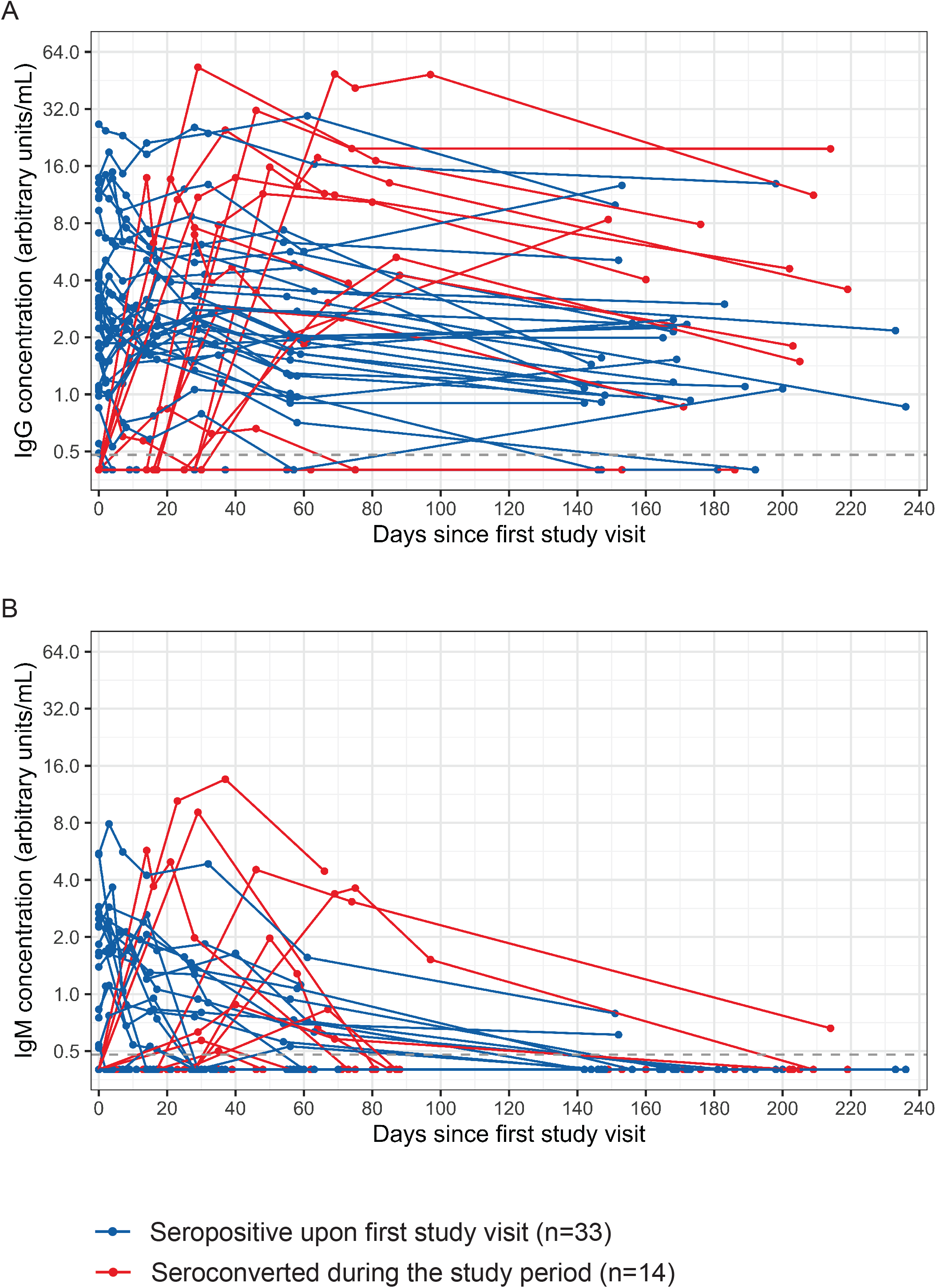
Antibody kinetics in 47 health care workers following SARS-CoV-2 infection (n=300 samples). IgG (A) and IgM (B) antibodies to the SARS-CoV-2 S-RBD are shown in health care workers who possessed IgG antibodies upon first study visit (n=33) or seroconverted during the study period (n=14). Lines connect samples collected from one individual.

### Correlates of protection against SARS-CoV-2 infection and disease severity

SARS-CoV-2 circulated at low levels in Philadelphia during the summer of 2020 but infections increased during the fall of 2020 and the subsequent winter (**Figure 1A**). We invited all participants to complete an online survey in January 2021 to report SARS-CoV-2 infections since the last blood draw, and over half of the participants (1,159) completed this survey. For purposes of this study, we defined infections that occurred during our initial spring/summer sampling period as ‘viral period one’ (infections that occurred before July 2, 2020) and infections that occurred after our initial sampling period as ‘viral period two’ (infections that occurred after July 2, 2020) (**Figure 1A**). We analyzed the last spring/summer serum sample collected from participants who were either infected or not infected during the second viral period to identify specific types of antibodies that were correlated with protection.

Forty-four of the 55 health care workers who were SARS-CoV-2 S-RBD seropositive during the first wave of SARS-CoV-2 infections responded to our January 2021 survey. Of these 44 participants who had detectable antibodies to the SARS-CoV-2 S-RBD, 1 seropositive participant (2.3%) reported a PCR-confirmed infection with SARS-CoV-2 during the second viral period in the fall of 2020. This participant experienced COVID-19 symptoms including cough and difficulty breathing. This participant entered our study as SARS-CoV-2 seropositive and SARS-CoV-2 PCR negative, and therefore we could not confirm that this participant had a SARS-CoV-2 infection during the first wave of virus circulation during the spring of 2020. It is possible that this individual was not previously infected with SARS-CoV-2 but instead possessed pre-pandemic cross-reactive S-RBD antibodies, which we have found were present in approximately 0.9% of individuals before the COVID-19 pandemic began (3). It is also possible that this individual was SARS-CoV-2 infected in the spring of 2020 and then re-infected with an antigenically distinct strain of SARS-CoV-2 in the fall of 2020, although we could not investigate this possibility since we were unable to obtain NP samples from the fall infection for sequencing.

Of the 1,115 health care workers who did not have detectable SARS-CoV-2 spike-RBD antibodies during the spring and summer of 2020 and who responded to our January 2021 online survey, 68 participants (6.1%) reported a lab-confirmed SARS-CoV-2 infection after the last blood draw, including 64 symptomatic infections. Two participants were hospitalized during the fall and winter because of COVID-19 (**Supplemental Table 2**). We completed additional serological assays using samples collected during the spring and summer of 2020 from the 68 SARS-CoV-2 spike-RBD seronegative individuals who were PCR-confirmed infected during the second viral period and 68 participants matched by age and sex who did not report SARS-CoV-2 infections after the last blood draw. This allowed us to evaluate correlates of protection associated with preventing SARS-CoV-2 infections in individuals who have not been previously exposed to the virus. We used ELISA to quantify antibodies against the SARS-CoV-2 full length spike (S-FL) protein and N protein, as well as antibodies against S-FL proteins from CCVs. Consistent with our previous study (3), we found that pre-infection antibodies reactive to SARS-CoV-2 S-FL protein and N protein were rare and at similar levels in health care workers who were and were not infected with SARS-CoV-2 during the second viral period (**Figure 3A**). Similarly, we found that antibody titers to S-FL from CCVs were not associated with protection from PCR-confirmed SARS-CoV-2 infections (**Figure 3A**).

**Figure 3.**
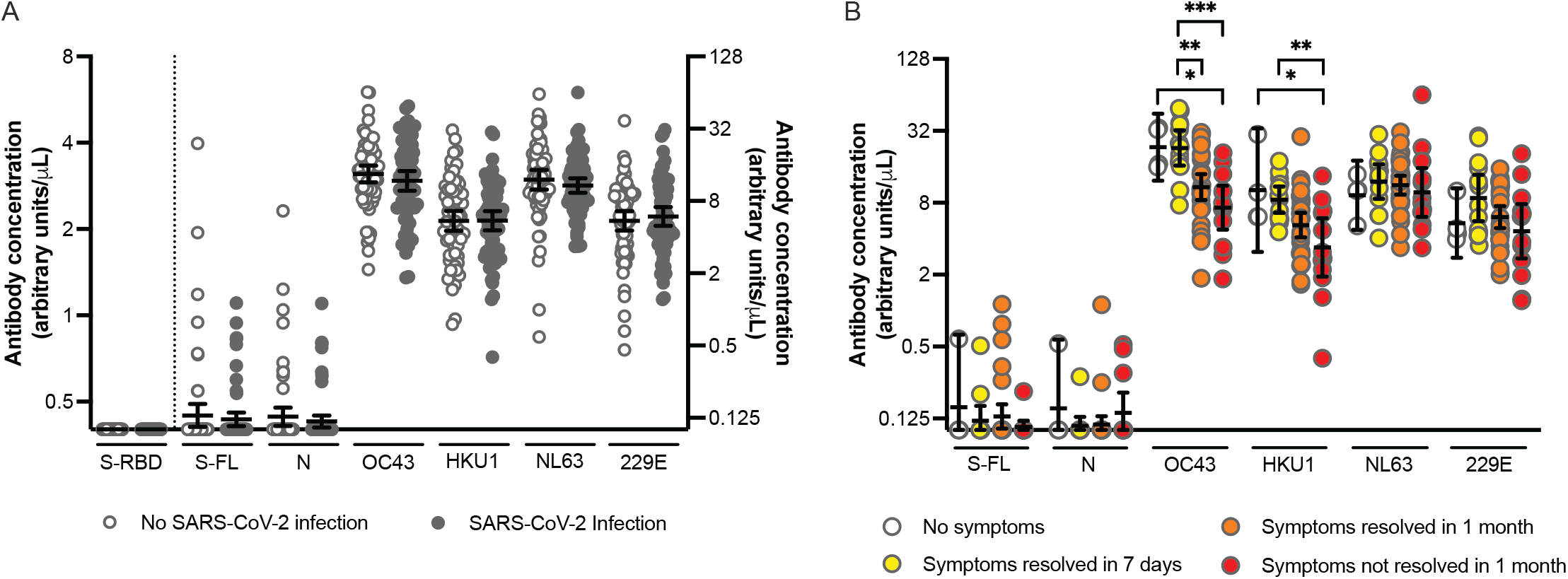
Correlation between pre-existing antibody concentrations and reported SARS-CoV-2 infections and duration of COVID-19 symptoms. (A) Pre-existing antibody concentrations in health care workers with (n=68) and without (n=68) SARS-CoV-2 infection after last blood draw. The control group without SARS-CoV-2 infection after last blood draw was matched to the infection group based on age and sex. Antibody concentrations were similar between infected and uninfected individuals (p>0.28 in unpaired t-tests using log_2_-transformed antibody concentrations). Antibody concentrations specific to S-RBD are projected on the left Y-axis and values below the cutoff (0.48) are set at 0.40. All other antibody concentrations are projected on the right Y-axis. And values below the limit of detection (0.20) are set at 0.10. Horizontal lines show the geometric mean concentrations and 95% confidence intervals. (B) Pre-existing antibody concentrations in health care workers who reported a PCR-confirmed SARS-CoV-2 infection and had no symptoms (n=4) or indicated symptom duration via an online survey (n=58). Symptom duration was grouped symptoms resolved within 7 days (n=13), symptoms resolved within 1 month (n=32) and symptoms not resolved within 1 month (n=13). Significant p-values (<0.05) are indicated above the graph (one-way ANOVA using log_2_-transformed antibody concentrations). Horizontal lines show the geometric mean concentrations and 95% confidence intervals. * p<0.05; ** p<0.01; *** p<0.001.

Next, we compared the relationship of pre-infection antibody levels with disease severity following SARS-CoV-2 infection using samples collected from SARS-CoV-2 S-RBD seronegative individuals who became infected during the second viral period. Our analysis included samples from 4 health care workers who reported asymptomatic SARS-CoV-2 infections and 58 participants who reported symptomatic SARS-CoV-2 infections (6 participants did not include information about symptoms and were therefore not included in the analyses). We found no correlation between symptom duration and pre-infection antibody levels reactive to SARS-CoV-2 S-FL and N proteins (**Figure 3B**). In contrast, we found a strong negative association of symptom duration and pre-infection antibody titers to OC43 and HKU1 S-FL proteins (**Figure 3B**). After adjusting via multivariate regression for age and sex, individuals with higher OC43 and HKU1 S-FL antibody titers had significantly fewer symptomatic days following SARS-CoV-2 infection (p=0.004 and p=0.030, respectively; **Supplemental Table 3**). There was no correlation between symptom duration and antibody titers to 229E and NL63 S-FL proteins (**Figure 3B**). This is interesting since 229E and NL63 are both alphacoronaviruses, whereas SARS-CoV-2, OC43, and HKU1 are all betacoronaviruses (βCoVs). While we found significant associations between OC43 and HKU1 antibody titers and SARS-CoV-2 symptom duration, we found few associations with pre-infection antibody titers and specific symptoms (**Supplemental Figure**). Since antibodies to CCVs are elevated after CCV infection and then slowly decline over time (6), individuals with higher OC43 and HKU1 antibody titers in our cohort were more likely recently infected with these common βCoVs. The mechanism underlying the apparent transient cross-protection between common βCoV infections and SARS-CoV-2 symptom duration in our cohort is unknown, but potentially involves cellular immunity since we found no association with pre-existing SARS-CoV-2-reactive antibodies and symptom duration following SARS-CoV-2 infections.

## Discussion

In this study we show that SARS-CoV-2 infections among health care workers at the University of Pennsylvania are relatively uncommon. Our study, along with others (14, 15) suggest that PPE and other precautions have efficiently limited the spread of SARS-CoV-2 within our hospitals. Consistent with other studies (16-19), we show that antibody responses elicited by SARS-CoV-2 are long-lived and detectable up to 140 days following infection in the majority of individuals. We identified one individual in our study who was potentially infected twice with SARS-CoV-2 but it is unclear if this individual was truly infected during the first wave of SARS-CoV-2 in Philadelphia. This individual was SARS-CoV-2 S-RBD antibody positive entering the study in the spring of 2020 and it is possible that this participant was not infected during the first SARS-CoV-2 wave but instead possessed pre-pandemic cross-reactive SARS-CoV-2 S-RBD antibodies that were present in approximately 0.9% of individuals before the pandemic began (3).

The primary goals of our study were initially to use serology to monitor asymptomatic and symptomatic infections of health care workers during the spring/summer of 2020 and to kinetically measure antibody levels after infection. Since SARS-CoV-2 continued circulating after the first wave in Philadelphia, we completed additional analyses to determine if antibody levels in serum samples collected in the summer of 2020 were associated with protection from subsequent SARS-CoV-2 infections. Consistent with our recent study (3), we found that pre-infection SARS-CoV-2 S-FL and N antibody levels were not associated with protection from SARS-CoV-2 infection during the fall and winter in individuals who were not infected with SARS-CoV-2 during the first viral wave. This is consistent with the observation that most pre-pandemic cross-reactive SARS-CoV-2 antibodies are non-neutralizing (3). Similarly, we found that pre-infection OC43, HKU1, 229E, and NL63 S-FL antibody titers were not associated with protection from SARS-CoV-2 infection.

Somewhat paradoxically, we found significant negative correlations between pre-infection OC43 and HKU1 antibody titers and SARS-CoV-2 symptom duration but we did not find correlations between pre-infection SARS-CoV-2 antibodies and SARS-CoV-2 symptom duration among individuals infected with SARS-CoV-2 for the first time during the second viral wave. Individuals with higher OC43 and HKU1 antibody titers experienced shorter duration of symptoms following SARS-CoV-2 infection. This apparent cross-protection appears to be specific to βCoV immunity since we found that antibody titers to the HKU1 and 229E alphacoronaviruses were not associated with reducing SARS-CoV-2 symptom duration. It may seem contradictory that OC43 and HKU1 antibody levels but not SARS-CoV-2 antibody levels are associated with reduced symptom duration in individuals who are infected with SARS-CoV-2 for the first time. However, the cross-protection afforded by common βCoVs is likely not mediated by rare antibodies that cross-react to SARS-CoV-2 proteins. Instead, this protection might be mediated by cellular immune responses, which can target epitopes that are conserved among common βCoVs and SARS-CoV-2 (20). Individuals who were more recently infected with common βCoVs have higher levels of antibodies against these viruses (6), and therefore elevated levels of antibodies against OC43 and HKU1 may serve as an ‘immunological stamp’ that dates how recently an individual was exposed to common βCoVs. Additional studies need to be completed to determine the temporal relationship between recent βCoV infections and reduced symptom duration following SARS-CoV-2 infections; however, our data are consistent with a recent electronic health record study that suggested that recent common coronavirus infections were associated with reducing the severity of COVID-19 (2). It is possible that T cells stimulated from recent βCoV infections (21) are involved with clearing virus and reducing symptom duration following SARS-CoV-2 infections. It is also possible that recent βCoV infections stimulate rare B cells that are quickly recalled following SARS-CoV-2 exposures.

Moving forward, it is possible that antibody titers to OC43 and HKU1 might be useful for predicting relative infection risk among individuals who have not yet encountered SARS-CoV-2. This type of information might be important for prioritizing vaccinations while the vaccine supply remains limited. One ironic implication of our study is that individuals who have efficiently socially distanced over the past year are potentially at higher risk of more severe SARS-CoV-2 symptoms since it is unlikely that these individuals have been recently infected with common βCoVs during social isolation. Additional studies will be required to fully understand the complex relationship between CCV immunity and SARS-CoV-2 susceptibility and temporal relationships between viral infections and cross-protection.

## Methods

### Study population and data collection

Health care workers at 3 hospitals in the University of Pennsylvania health system (Hospital of the University of Pennsylvania, Penn Presbyterian Medical Center, and Pennsylvania Hospital) were recruited between April 13, 2020 to May 20, 2020. Only health care workers with direct contact with patients or who worked on units where patients with COVID-19 received care were included in this study. We excluded anyone who was previously diagnosed with a SARS-CoV-2 infection. Characteristics of the 2,043 health care workers in our study are reported in **Supplemental Table 1**. We collected serum samples from each participant and quantified SARS-CoV-2 S-RBD antibodies by ELISAs within 36-48 hours after sample collection. We collected NP swabs from all health care workers who possessed SARS-CoV-2 S-RBD IgG and/or IgM antibodies and we completed SARS-CoV-2 PCR testing on these samples to identify active or recent infections. Health care workers who were seronegative at baseline visit were invited for follow-up visits every 2 weeks until 2 July 2020 to identify active SARS-CoV-2 infections throughout the study period. In addition, we received serum samples from 8 health care workers who were seronegative at baseline visit and had a positive NP PCR test outside of our study. Seropositive health care workers were enrolled in a follow-up study to collect additional blood samples up to 236 days post seroconversion.

Participants filled out an online survey at time of enrollment to collect participant characteristics, including COVID-19 symptom information. A second online survey was sent to all participants in January 2021 to collect information on new SARS-CoV-2 infections that occurred after the last blood draw. Based in this information, additional SARS-CoV-2 and common coronavirus ELISAs were completed in a subset of participants to study pre-infection antibodies. Informed consent was collected from all participants prior to the baseline visit. This study was approved by the Institutional Review Board of the University of Pennsylvania under IRB #842847.

### Enzyme-linked immunosorbent assay (ELISA)

ELISAs measuring antibodies against SARS-CoV-2 and against OC43, HKU1, 229E and NL63 were completed as previously described (13). SARS-CoV-2 nucleocapsid (N) protein and OC43, HKU1, 229E, and NL63 S-FL proteins were purchased (Sino Biological). Plasmids encoding the SARS-CoV-2 S-FL and the S-RBD were provided by Florian Krammer (Mt. Sinai). SARS-CoV-2 S-FL and S-RBD were produced in 293F cells and purified using Ni-NTA (Qiagen). Each well in an ELISA plate (Immulon 4 HBX, Thermo Scientific) was coated with 50 μL PBS or recombinant protein (2 μg/mL SARS-CoV-2 antigen or 1.5 μg/mL OC43 antigen) and plates were incubated overnight at 4°C. Wells coated with only PBS were used to measure background signal for each sample. The next day, plates were washed with PBS containing 0.1% Tween-20 (PBS-T) and incubated for 1 hour with PBS-T supplemented with 3% non-fat milk powder and 0.1% Tween-20. Heat-inactivated serum samples were diluted in PBS-T supplemented with 1% non-fat milk powder and 0.1% Tween-20 (dilution buffer). ELISA plates were washed with PBS-T and 50 μL serum dilution was added to each well. After 2 hours of incubation, plates were washed with PBS-T and 50 μL of 1:5,000 diluted goat anti-human IgG-HRP (Jackson ImmunoResearch Laboratories) or 1:1,000 diluted goat anti-human IgM-HRP (SouthernBiotech) was added to each well. Plates were incubated for 1 hour and washed with PBS-T before 50 μL SureBlue TMB Substrate (KPL) was added to each well. After 5 minutes, the reaction was stopped by adding 25 μL of 250 mM hydrochloric acid. Plates were read at an optical density (OD) of 450 nm using the SpectraMax 190 microplate reader (Molecular Devices). Monoclonal antibody CR3022 (for SARS-CoV-2 S and RBD ELISA) or in-house created serum pool (for SARS-CoV-2 N ELISA and common coronavirus ELISAs) was included on each plate to convert OD values into relative antibody concentrations. Plasmids to express the CR3022 monoclonal antibody were provided by Ian Wilson (Scripps).

### Statistical analysis

Health care workers with serum IgG and/or IgM concentration above 0.48 μg/mL in SARS-CoV-2 S-RBD ELISAs were considered seropositive. This cutoff results in a pre-pandemic cross-reactive rate of 0.6% for IgG and 0.5% for IgM, as was described previously (13). SARS-CoV-2 S-RBD antibody concentrations below this cutoff at 0.48 arbitrary units/mL were assigned a value of 0.40 arbitrary units/mL. All other antibodies below the limit of detection (0.20 arbitrary units/mL) were assigned a value of 0.10 arbitrary units/mL. Antibody concentrations were log_2_-transformed for analysis and geometric mean concentrations with 95% confidence intervals were reported unless stated otherwise. Standard descriptive analyses were used as appropriate including the Chi-squared test, paired and unpaired *t* tests, Mann-Whitney test and one-way ANOVA with Bonferroni correction to adjust for multiple comparisons. Pre-existing antibody titers were fitted to symptom durations in days in separate linear models via logistic regression with a logit link function, adjusting for age and sex. Statistical significance was set at p<0.05. Prism version 9 (GraphPad) and R version 3.5.3 were used for analyses.

## Data Availability

All data are included in the manuscript.

## ACKNOWLEDGEMENTS

This work was supported by institutional funds from the University of Pennsylvania and NIH U19AI082630 (EJW and SEH) and UM1AI069534 (IF). A.B. and J.W. were supported by the NIH award number UL1TR001878. EMA was supported by the NIH Training in Virology T32 Program (T32AI007324) and CPA was supported by the NIH Emerging Infectious Diseases T32 Program (T32AI055400). The content is solely the responsibility of the authors and does not necessarily represent the official views of the NIH. We thank J. Lurie, J. Embiid, J. Harris, and D. Blitzer and J. and M. Greenberg for philanthropic support. We would like to thank the UPenn COVID Processing Unit, who are comprised of individuals from diverse laboratories at the University of Pennsylvania who volunteered time and effort to enable study of COVID-19 patients during the pandemic: Sharon Adamski, Zahidul Alam, Mary M. Addison, Katelyn T. Byrne, Aditi Chandra, Kurt D’Andrea, Hélène C. Descamps, Nicholas Han, Yaroslav Kaminskiy, Shane C. Kammerman, Justin Kim, Allison R. Greenplate, Jacob T. Hamilton, Nune Markosyan, Julia Han Noll, Dalia K. Omran, Ajinkya Pattekar, Eric Perkey, Elizabeth M. Prager, Dana Pueschl, Austin Rennels, Jennifer B. Shah, Jake S. Shilan, Nils Wilhausen, Ashley N. Vanderbeck. We thank F. Krammer (Mt. Sinai) for sending us the SARS-CoV-2 spike RBD expression plasmids in the early days of the COVID-19 pandemic, which was critical for initiating our health care worker cohort study during the spring of 2020.

## Figure legend

**Supplemental Figure.**
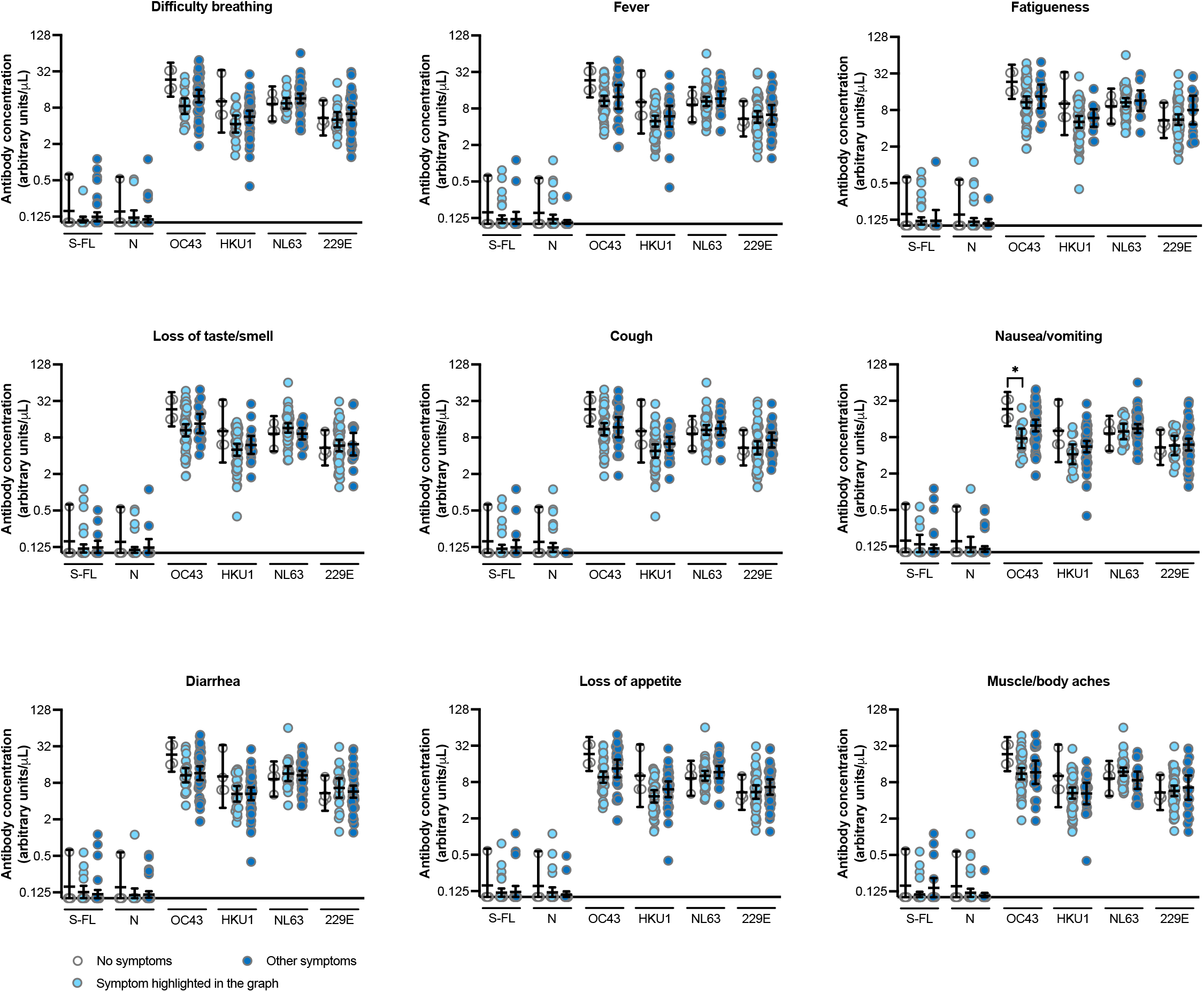
Correlation between pre-existing antibody concentrations and reported COVID-19-related symptoms in symptomatic (n=64) and asymptomatic (n=4) health care workers with a PCR-confirmed SARS-CoV-2 infection. Significant p-values (<0.05) are indicated above the graph (one-way ANOVA using log_2_-transformed antibody concentrations). Horizontal lines show the geometric mean concentrations and 95% confidence intervals. * p<0.05.

**Supplemental Table 1.**
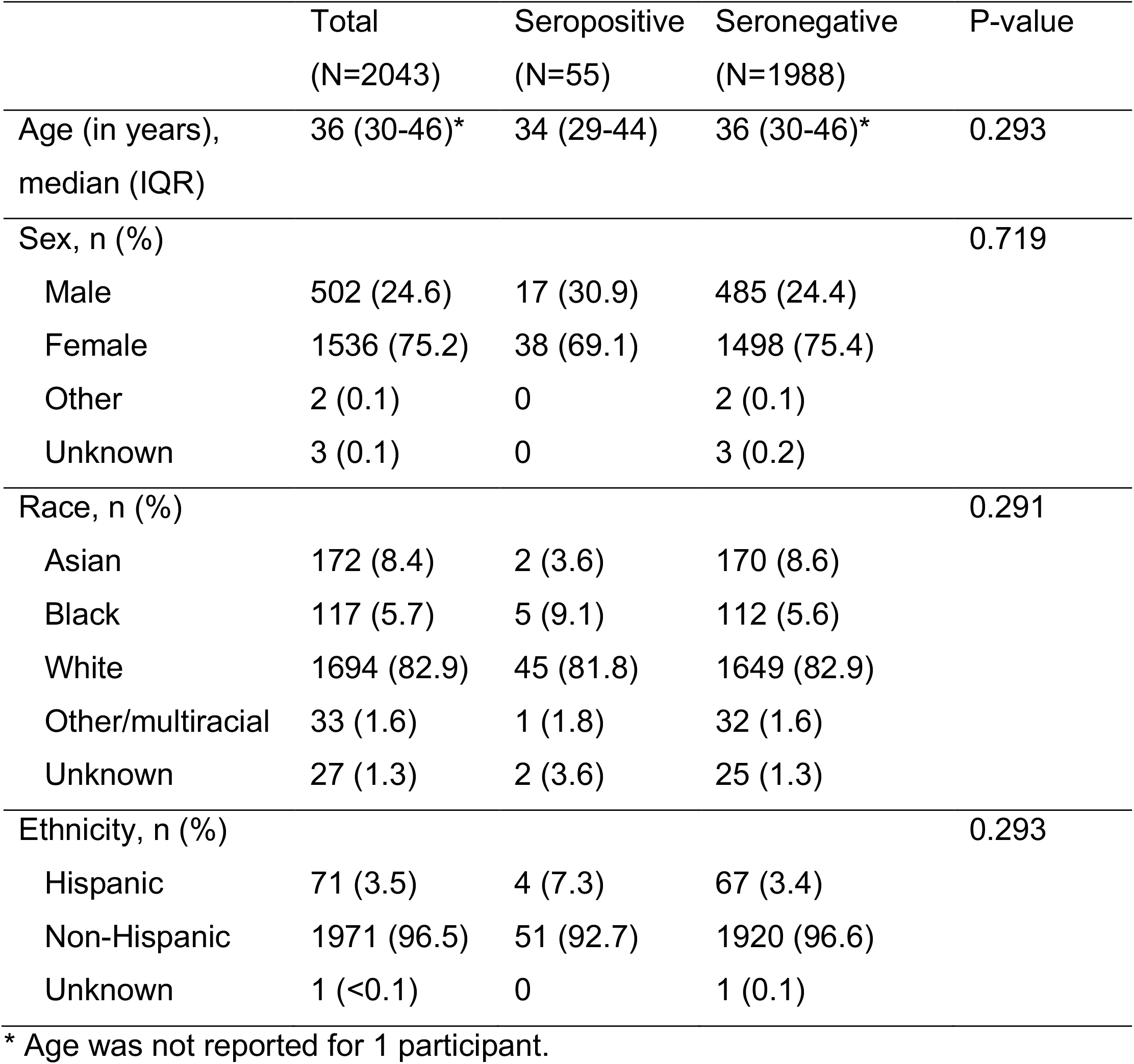
Participant characteristics.

**Supplemental Table 2.** Characteristics of 64 participants who had no detectable S-RBD antibodies during the spring and summer of 2020 and reported via the online survey that they had a symptomatic lab-confirmed SARS-CoV-2 infection after the last blood draw.

|  | **Total (N=64)** |
| --- | --- |
| Symptoms, n (%) |  |
| Cough | 42 (65.6) |
| Fever or chills | 42 (65.6) |
| Nausea/vomiting | 13 (20.3) |
| Diarrhea | 20 (31.3) |
| Unusual fatigue | 51 (79.7) |
| Loss of taste or smell | 47 (73.4) |
| Difficulty breathing | 17 (26.6) |
| Loss of appetite | 34 (53.1) |
| Muscle/body aches | 45 (70.3) |
| Other | 23 (35.9) |
| Estimated date of SARS-CoV-2 infection, n (%) |  |
| July 2020 | 6 (9.4) |
| August 2020 | 1 (1.6) |
| September 2020 | 0 |
| October 2020 | 9 (14.1) |
| November 2020 | 14 (21.9) |
| December 2020 | 30 (46.9) |
| January 2021 | 4 (6.3) |
| Resolution of symptoms, n (%) |  |
| Within 7 days | 13 (20.3) |
| Within 1 month | 32 (50.0) |
| More than 1 month | 13 (20.3) |
| Unknown | 6 (9.4) |
| Hospitalization because of COVID-19, n (%) | 2 (3.1) |

**Supplemental Table 3.** Effects of log_2_ pre-existing antibody titers, age and sex on symptom duration (days) via multivariate regression.

|  | **Estimate** | **SE** | **P-value** |
| --- | --- | --- | --- |
| S-FL | -2.731 | 2.685 | 0.313 |
| Age | 3.204 | 2.697 | 0.240 |
| Sex (male) | 5.675 | 7.964 | 0.479 |
| N | 1.895 | 2.698 | 0.485 |
| Age | 3.606 | 2.712 | 0.189 |
| Sex (male) | 6.299 | 7.997 | 0.434 |
| OC43 | -8.166 | 2.739 | 0.004 |
| Age | 0.170 | 2.752 | 0.951 |
| Sex (male) | 4.226 | 7.498 | 0.575 |
| HKU1 | -5.867 | 2.630 | 0.030 |
| Age | 2.392 | 2.644 | 0.369 |
| Sex (male) | 5.559 | 7.704 | 0.474 |
| NL63 | -2.354 | 2.683 | 0.384 |
| Age | 3.548 | 2.697 | 0.194 |
| Sex (male) | 6.218 | 7.973 | 0.439 |
| 229E | -3.054 | 2.846 | 0.288 |
| Age | 2.840 | 2.742 | 0.305 |
| Sex (male) | 8.483 | 8.255 | 0.308 |

